# Peptidomic profiling reveals extracellular matrix remodeling signatures discriminative of multiple myeloma

**DOI:** 10.64898/2026.01.08.26343675

**Authors:** Maria Frantzi, Melika Ahangar, Antonia Vlahou, Harald Mischak, Irene Solia, Foteini Theodorakakou, Christine Ivy Liacos, Jerome Zoidakis, Evangelos Terpos, Meletios A. Dimopoulos, Efstathios Kastritis

## Abstract

Multiple myeloma (MM) evolves from monoclonal gammopathy of undetermined significance (MGUS) and smoldering MM (SMM) with annual progression rates of 1% and 10%, respectively. Current risk models don’t fully capture the underlying dynamic molecular processes. We hypothesized that urinary peptides reflect disease-specific microenvironmental alterations in plasma cell dyscrasias. To test this hypothesis, capillary electrophoresis–mass spectrometry CE–MS was applied to profile the urinary peptidome of 314 individuals, including a discovery group (42 MGUS, 27 SMM, 14 MM), an independent validation group (45 MGUS, 9 SMM, 7 MM, 9 with plasmacytoma), 86 without underlying malignancy, and 75 patients with impaired kidney function. 121 peptides were significantly altered between MM and MGUS and displayed a monotonic abundance trend across the MGUS–SMM–MM continuum. These peptides predominantly derived from collagens, beta-2 microglobulin, alpha-1 antitrypsin, and antithrombin-III. Integration of these 121 peptides into a support vector machine classifier achieved an area under the curve of 0.94 (0.85–0.99; 95% CI) in the independent validation cohort, with 100% sensitivity and 82% specificity for MM detection. The finding that urinary peptides enable non-invasive molecular discrimination of MM from precursor states represents a solid basis for a prospective evaluation in prognosis and detection of progression.

**Significance Statement:** Progression from monoclonal gammopathy of undetermined significance (MGUS) or smoldering myeloma (SMM) to active multiple myeloma (MM) remains difficult to predict in routine clinical practice. Current risk assessment based on the International Myeloma Working Group (IMWG) criteria primarily relies on clinical and biochemical variables. This study identifies myeloma-specific urinary peptide signatures reflecting extracellular matrix (ECM) remodeling. In a cohort of 314 patients, CE–MS–based urinary peptidomic analysis yielded a 121-peptide ECM-derived classifier that accurately differentiated active MM from precursor conditions, achieving 100% sensitivity and 82% specificity upon independent validation. Importantly, gradual changes in peptide abundance with disease evolution from MGUS to SMM to MM suggest that urinary ECM-related peptide fragments reflect stage-associated molecular changes across the MGUS–SMM–MM continuum. These findings represent a solid basis for the evaluation of the value of this classifier in predicting progression in a prospective study.

## Introduction

Multiple myeloma (MM) is a plasma-cell disorder characterized by clonal expansion of neoplastic plasma cells in the bone marrow and excessive production of monoclonal immunoglobulins, ultimately resulting in end-organ damage [1]. MM accounts for approximately 10% of hematologic malignancies and, despite substantial therapeutic advances, remains largely incurable [2]. MM develops through a well-defined biological continuum, beginning as monoclonal gammopathy of undetermined significance (MGUS) and progressing through asymptomatic (smoldering MM / SMM) to symptomatic disease [3]. MGUS affects approximately 3–5% of individuals over 50 years of age and progresses to MM at an average rate of 1% per year, whereas SMM can progress at rates approaching 10% annually or even higher in those with high-risk features [4]. Existing clinical risk models based on serum M-protein concentration, free light-chain ratio, and bone marrow plasma-cell infiltration provide useful but static stratification and do not reflect the biological processes underlying malignant transformation [3]. Increasing evidence indicates that progression from MGUS to MM is not solely driven by tumor-intrinsic mutations, but also by remodeling of the bone marrow microenvironment (BME), including extracellular matrix (ECM) degradation and immune–stromal interactions [5, 6]. Biomarkers capable of capturing these dynamic processes represent a major unmet clinical need.Malignant plasma cells rely on reciprocal interactions with stromal and endothelial cells, cytokines, immune regulators, and the ECM, creating a permissive niche that promotes tumor survival, immune escape, and drug resistance, especially within the bone marrow [7, 8]. Notably, ECM-derived peptide fragments are detectable systemically and have been associated with tumor burden and immune dysregulation [9, 10]. These findings highlight ECM turnover as a biologically relevant source of measurable circulating biomarkers. Conventional approaches for assessing ECM remodeling, such as bone marrow biopsy followed by immunohistochemistry and serological tests, have provided mechanistic insights but have significant limitations. Bone marrow biopsies are invasive, spatially heterogeneous, and unsuitable for frequent sampling, while tissue-based proteomics cannot distinguish local from systemic remodeling [11, 12]. These limitations are particularly impactful on precursor conditions (MGUS, SMM), where subtle microenvironmental changes may precede clinical progression by years, yet remain undetectable using current serological markers. Consequently, a non-invasive and sensitive, molecular-based assay to quantify ECM remodeling without the need for serial bone marrow biopsies can be of value.

Urinary peptidomics offers a promising solution. Unlike serum or tissue proteomics, the urinary peptidome comprises naturally occurring low-molecular-weight peptides that pass the glomerular filter, providing a molecular profiling of endogenous protein turnover while minimizing confounding effects from ex vivo processing. Capillary electrophoresis coupled to mass spectrometry (CE–MS) offers analytical robustness, precise peptide quantification, excellent inter-laboratory reproducibility, and is currently the only urinary proteomics technology with FDA-compliant quality-control pipelines [13, 14].

CE–MS urinary classifiers reflect complex biological processes, outperforming single-analyte biomarkers, and have been developed for multiple contexts of use in chronic kidney disease, cardiovascular disease, and several malignancies [15, 16]. Urinary peptides can also be assessed repeatedly, at low procedural risk, making them well-suited for longitudinal surveillance. Importantly, CE-MS technology has proven to be particularly effective in resolving urinary peptides of ECM origin, paving the way for its application in plasma cell neoplasia.

Herein, we hypothesized that proteolytic fragments derived from ECM components and immune-regulatory proteins are released into circulation during MM initiation and progression and ultimately excreted in urine, where they may serve as quantifiable biomarkers specific to MM. To test this hypothesis, in this feasibility cross-sectional study, we performed CE–MS to profile and compare the urinary peptidomes between patients with MGUS, SMM, and MM, intending to determine whether systemic ECM remodeling is detectable in urine from patients with MM and/or other precursor conditions, and whether urinary peptide patterns can discriminate precursor conditions from active MM. A graphical overview of the study concept and analytical workflow is presented in **Figure 1**.

**Figure 1:**
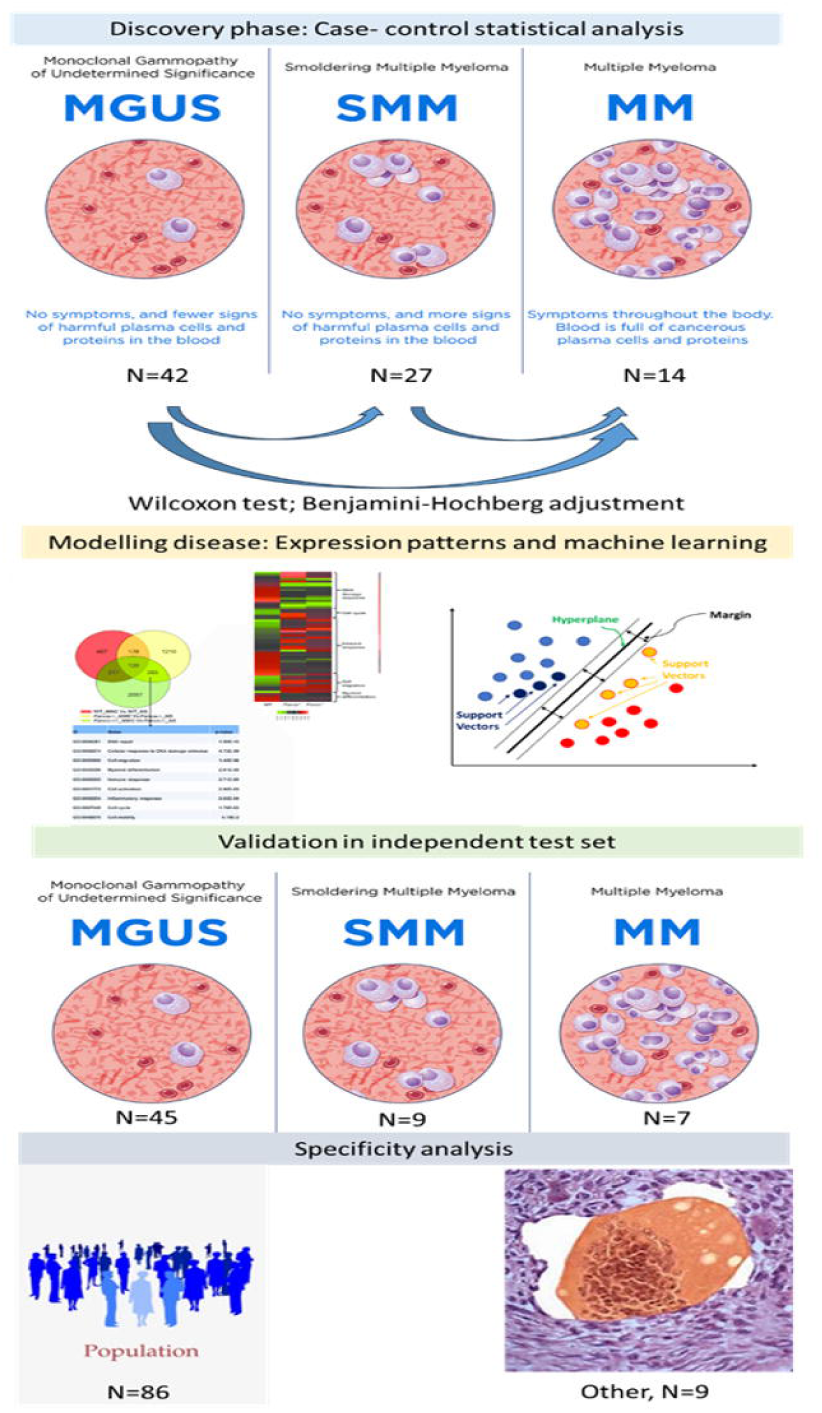
Graphical overview of the study design, including a discovery and a validation phase, along with a preliminary specificity analysis in populational cohorts and a closely related malignancy (plasmacytoma).

## Materials and Methods

### Study Design and Patient Cohorts

This cross-sectional study was conducted within the Horizon Europe project ELMUMY (Grant Agreement No. 101097094) and aimed to identify urinary peptide signatures specific to MM. A total of 153 urinary peptide profiles from patients within ELMUMY were analyzed by CE–MS in a two-phase study design consisting of a discovery and an independent validation cohort, along with a disease-specificity assessment. The discovery cohort consisted of 83 patients (42 MGUS, 27 smoldering MM, 14 symptomatic MM). An independent validation cohort included 61 subjects (45 MGUS, 9 SMM, 7 MM). In addition, nine urinary profiles from patients with related plasma-cell disorders (plasmacytoma), as per availability, were incorporated for biological specificity assessment. The CE–MS datasets included in this analysis fulfilled established MSI-compliant quality criteria, including peptide signal normalization and migration-time calibration, as described previously [13]. All MGUS, SMM, and MM samples were obtained from patients visiting the Hematology/Oncology Unit of the Department of Clinical Therapeutics, National and Kapodistrian University of Athens. The diagnosis of MGUS, SMM, and MM was based on the IMWG 2014 criteria[17]. The sample collection was made under institutional review board approval (IRB protocol number 111368/ 50/2022) in accordance with the Declaration of Helsinki and General Data Protection Regulation. Written informed consent was obtained from all subjects prior to participation. Summary descriptives of the patient’s demographic and biochemical variables are provided in **Table 1**. Estimated glomerular filtration rate (eGFR) was calculated using the CKD-EPI (Chronic Kidney Disease Epidemiology Collaboration) equation as follows: eGFR = 142 × min (Scr/K, 1)^α × max (Scr/K, 1)^−1.200 × 0.9938^age × 1.012 (if female), where Scr is serum creatinine (mg/dL), K is 0.7 for females and 0.9 for males, and α is −0.241 for females and −0.302 for males.[18] To assess the specificity of the peptide signatures, 86 urinary peptide datasets from healthy individuals were retrieved from the Mosaiques Diagnostics Urinary Peptidomics Database and served as non-malignant controls (reference cohort)[19]. Additionally, CE-MS data from urine samples from 75 patients at risk or diagnosed with impaired kidney function (25 individuals at risk for developing acute kidney injury/ AKI, 25 patients diagnosed with stage 3 AKI, and 25 patients diagnosed with chronic kidney disease / CKD) were investigated to assess the potential confounding effects of decreased renal function that is present within MM patients[20].

**Table 1.** Cohort Demographics and Clinical Parameters

| Discovery group |  |  |  |
| --- | --- | --- | --- |
| Clinical/ biochemical variable | MGUS (n=42) | MM (n=14) | SMM (n=27) |
| BMI (mean ± SD) | 26.51 ± 5.05 | 28.76 ± 5.58 | 25.97 ± 3.84 |
| eGFR (mean ± SD) | 80.13 ± 25.64 | 61.98 ± 35.32 | 83.84 ± 25.93 |
| Age (mean ± SD) | 64.86 ± 13.41 | 72.57 ± 7.10 | 69.70 ± 11.26 |
| Gender | 28.57% male | 35.71% male | 55.56% male |
| Nr. Peptides (mean ± SD) | 4202.43 ± 1259.58 | 4459.57 ± 1341.23 | 4172.59 ± 1265.94 |
| Validation group |  |  |  |
| Clinical/ biochemical variable | MGUS (n=45) | MM (n=7) | SMM (n=9) |
| BMI (mean ± SD) | 26.54 ± 4.44 | 28.83 ± 4.20 | 26.30 ± 4.63 |
| eGFR (mean ± SD) | 66.48 ± 26.94 | 78.70 ± 25.87 | 75.75 ± 22.09 |
| Age (mean ± SD) | 70.33 ± 11.97 | 72.45 ± 10.62 | 68.95 ± 13.13 |
| Gender | 52.38% male | 33.33% male | 66.67% male |
| Nr. Peptides (mean ± SD) | 2158.67 ± 658.59 | 2554.43 ± 806.77 | 1752.22 ± 372.37 |
| Variable in discovery | MGUS vs SMM | MGUS vs MM | SMM vs MM |
| AGE | 0.1863 | 0.0597 | 0.4936 |
| BMI | 0.6029 | 0.2217 | 0.1756 |
| eGFR | 0.4672 | 0.0223 | 0.0400 |
| Variable in validation | MGUS vs SMM | MGUS vs MM | SMM vs MM |
| AGE | 0.6699 | 0.8053 | 0.9048 |
| BMI | 0.7929 | 0.3538 | 0.5169 |
| eGFR | 0.3758 | 0.3538 | 1.0000 |
|  | Overall p (Kruskal–Wallis) |  |  |
| AGE | 0.8721 |  |  |
| BMI | 0.5826 |  |  |
| eGFR | 0.4529 |  |  |

### Urine sampling, Sample Preparation and Peptide Extraction

The second urine of the morning was collected and immediately frozen and stored at <-20°C. Urine samples were processed according to the standardized CE–MS workflow established by Mosaiques Diagnostics to ensure inter-study comparability and analytical reproducibility [13]. Briefly, 700 µL of thawed urine was mixed 1:1 with alkaline buffer (2 M urea, 10 mM NHLOH, 0.02% SDS; pH 10.5) to denature proteins and stabilize endogenous urinary peptides. The mixture was centrifuged through 20 kDa molecular weight cut-off filters (Centrisart, Sartorius, Göttingen, Germany) to remove high-molecular-weight proteins and cellular debris. The filtrate (1.1 mL) was desalted using PD-10 columns (GE Healthcare, Munich, Germany) pre-equilibrated with 0.01% NHLOH. After washing (1.9 mL) and elution (2.0 mL of HPLC-grade water), samples were lyophilized and stored at –80 °C until CE–MS analysis.

### Capillary Electrophoresis–Mass Spectrometry (CE–MS)

Urinary peptide profiling was performed on a P/ACE MDQ capillary electrophoresis system (Beckman Coulter, Fullerton, CA, USA) coupled via a grounded electrospray interface (Agilent Technologies) to a Micro-TOF mass spectrometer (Bruker Daltonics, Bremen, Germany). Lyophilized samples were reconstituted in HPLC-grade water, and 250 nL were injected hydrodynamically (2.0 psi, 99 s). Peptide separation was conducted under reverse polarity at 25 kV for 30 min, followed by a pressure-driven elution phase (0.5 psi, 34 min). Mass spectra were acquired every 3 seconds over an m/z range of 350–3000. Mass accuracy and migration time were calibrated using internal polypeptide standards .Peptide ion signal intensities were calibrated and normalized using 29 endogenous collagen fragments as internal standards, following the established MOS quality-control pipeline.

### Data Processing and Peptide Feature Extraction

Raw CE–MS data were processed using MosaiquesVisu and MosaFinder software (Version 6.2, Mosaiques Diagnostics GmbH, Hanover, Germany). Molecular features were extracted based on molecular mass, normalized migration time, and signal intensity. Mass accuracy and migration time calibration were performed using internal polypeptide standards. Spectral ion intensities were normalized using 29 consistently detected endogenous collagen fragments, as described previously [13]. Peptides detected in at least 30% of samples within any clinical group were retained for further analysis. Signal deconvolution, isotopic clustering, and molecular alignment followed MSI-compliant CE–MS criteria [21]. Peptide intensities were log-transformed before statistical and machine learning analysis. Feature retention statistics are provided in **Table 1**. Analytical performance evaluation followed FDA bioanalytical validation principles adapted for urinary peptide profiling. Instrument stability was monitored by repeated measurement of a pooled urine reference across analytical batches. Seven synthetic calibrant peptides were spiked at fixed concentrations to assess mass accuracy and signal linearity. Quality-control (QC) samples consisting of pooled urine from healthy donors were analyzed twice daily over multiple analytical days. Selectivity, linearity, and system suitability were confirmed according to established protocols [22].

#### Heatmap Generation and Peptide Expression Normalization

Heatmap plots were generated using the normalized mean abundance of the peptides across the three disease groups (MGUS, SMM, and MM; **Supplementary Table S1**). Mean peptide abundances for each group were extracted. For each peptide, a row-wise Z-score normalization was applied to highlight relative up- or down-regulation patterns independent of absolute intensity differences. Peptides were ordered according to their fold change between MM and MGUS (FC MM/MGUS) to enhance visualization of progressive expression trends. Heatmaps were visualized using a diverging color scale centered at zero (Z = 0), where positive values indicate relative up-regulation and negative values indicate down-regulation.

### Peptide Sequencing and Protein Identification

Selected urinary peptides were sequenced by tandem mass spectrometry (MS/MS) using a Q Exactive instrument (Thermo Fisher Scientific, Bremen, Germany) operated in Data Dependent Acquisition mode coupled to a P/ACE MDQ capillary electrophoresis system (Beckman Coulter, Fullerton, CA, USA). Peptide identification was performed in Proteome Discoverer (version 2.4) against the Swiss-Prot human proteome database, based on accurate mass, CE migration time, and charge-state matching. Only identifications with a false-discovery rate (FDR) < 1% were accepted. Representative MS/MS spectra were manually reviewed to confirm sequence quality.

### Statistical Analysis

Peptide intensities were log-transformed before the statistical analysis within the discovery patient groups. Group comparisons (MGUS vs SMM vs MM) were performed using the Mann–Whitney U test, followed by Benjamini–Hochberg correction for multiple testing within the discovery patient groups. Peptides with adjusted p-value < 0.05 were considered statistically significant. A peptide was considered monotonic if it was BH-significant in the MGUS vs MM comparison and exhibited a consistent direction of change in the MGUS vs SMM and SMM vs MM contrasts. Receiver operating characteristic (ROC) curves and area under the curve (AUC) values were calculated to evaluate discriminatory performance.

### Machine Learning Classification

A urinary peptide-based classifier was developed using support vector machine (SVM) modeling implemented in MosaCluster (Version 1.6.5, Mosaiques Diagnostics, Hanover, Germany). Feature input consisted of all log-transformed peptide intensities from those that were considered statistically significant in the discovery set, after Benjamini–Hochberg correction for multiple testing. The classifier was constructed using discriminatory peptides differentiating MGUS from MM in the discovery set. Model training was performed using an SVM with a radial basis function (RBF) kernel in the discovery set. Hyperparameters were optimized by five-fold stratified cross-validation. Classifier performance was evaluated within the discovery cohort using cross-validation and subsequently tested in the independent validation set. Feature selection following statistical analysis and SVM model optimization were performed exclusively within the discovery cohort, and the validation cohort was not used at any stage of model training or parameter tuning. Validation was performed in the independent patient groups using MosaDiagnostics (version 1.2.6, Mosaiques Diagnostics, Hanover, Germany). The performance of the SVM-based classifier was evaluated based on the pre-defined cut-off that was estimated in the discovery set. ROC plots were generated, and the respective confidence intervals (CI) were calculated using MedCalc statistical software (Version 12.7.5.0 ). The DeLong test was then used to compare the values obtained. The classification scores between the patient groups were compared using the Kruskal-Wallis rank sum test in MedCalc (Version 12.7.5.0).

### Pathway and Protein–Protein Interaction (PPI) Analysis

Proteins corresponding to the classifier peptides were annotated and functionally enriched using Metascape (www.metascape.org), employing Gene Ontology Biological Processes (GO-BP) and Reactome pathway databases as background. Enrichment was considered significant for p < 0.05, enrichment factor > 1.5, and minimum pathway membership ≥ 3 genes. Protein–protein interaction (PPI) networks were generated using STRING (Version 12) with a high-confidence interaction score ≥ 0.70 and visualized in Cytoscape (Version 3.9). Hub nodes were identified based on degree and betweenness centrality.

## Results

### Study Cohorts and Clinical Characteristics

A total of 314 urinary CE–MS peptide profiles were analyzed, representing a MM discovery group (83 MGUS, SMM, MM patients), a validation patient group (61 MGUS, SMM, MM patients, and 9 patients with plasmacytoma, which isa closely related malignancy), a reference cohort (86 individuals without underlying malignancy), and 75 patients at risk of or with impaired renal function. No significant differences were observed across MGUS, SMM, and MM groups for age or BMI (all p > 0.05). Overall renal function, assessed by estimated glomerular filtration rate (eGFR) calculated using the CKD-EPI equation, was comparable across MGUS, SMM, and MM groups. However, within the discovery cohort, renal function was significantly impaired in patients with MM compared to those with MGUS and SMM, likely reflecting the presence of kidney injury, as summarized in **Table 1**.

### Differential peptide analysis revealed 121 peptides with monotonic abundance trends across disease stages

After peptide filtering at a ≥30% detection threshold, differential abundance analysis revealed the largest molecular separation between MGUS and MM, followed by SMM versus MM. In total, 121 peptides were significant after BH correction (adjusted p < 0.05) in MGUS versus MM comparisons. These peptides showed robust discriminatory performance, with individual AUC values ranging between 0.70 and 0.95. In the SMM versus MM comparison, only three, all including collagen fragments, were found significant after adjustment for multiple testing. By contrast, MGUS versus SMM yielded no significantly changed peptides, consistent with a continuous biological transition between precursor stages. The 121 significant peptides were defined as myeloma-associated biomarkers, being significantly altered between patients with MM and MGUS, and subjected to downstream modeling. Several of these exhibited monotonic directional changes across the MGUS–SMM–MM disease continuum, including increasing urinary excretion of βL-microglobulin, increased serum levels of which are considered as a marker for high tumor burden and worse prognosis of MM[23]. The overlap between significant peptides (based on nominal p value) across pairwise comparisons is illustrated in **Figure 2A**, while monotonic abundance trajectories for the 121 progression-associated peptides are shown in **Figure 2B**.

**Figure 2.**
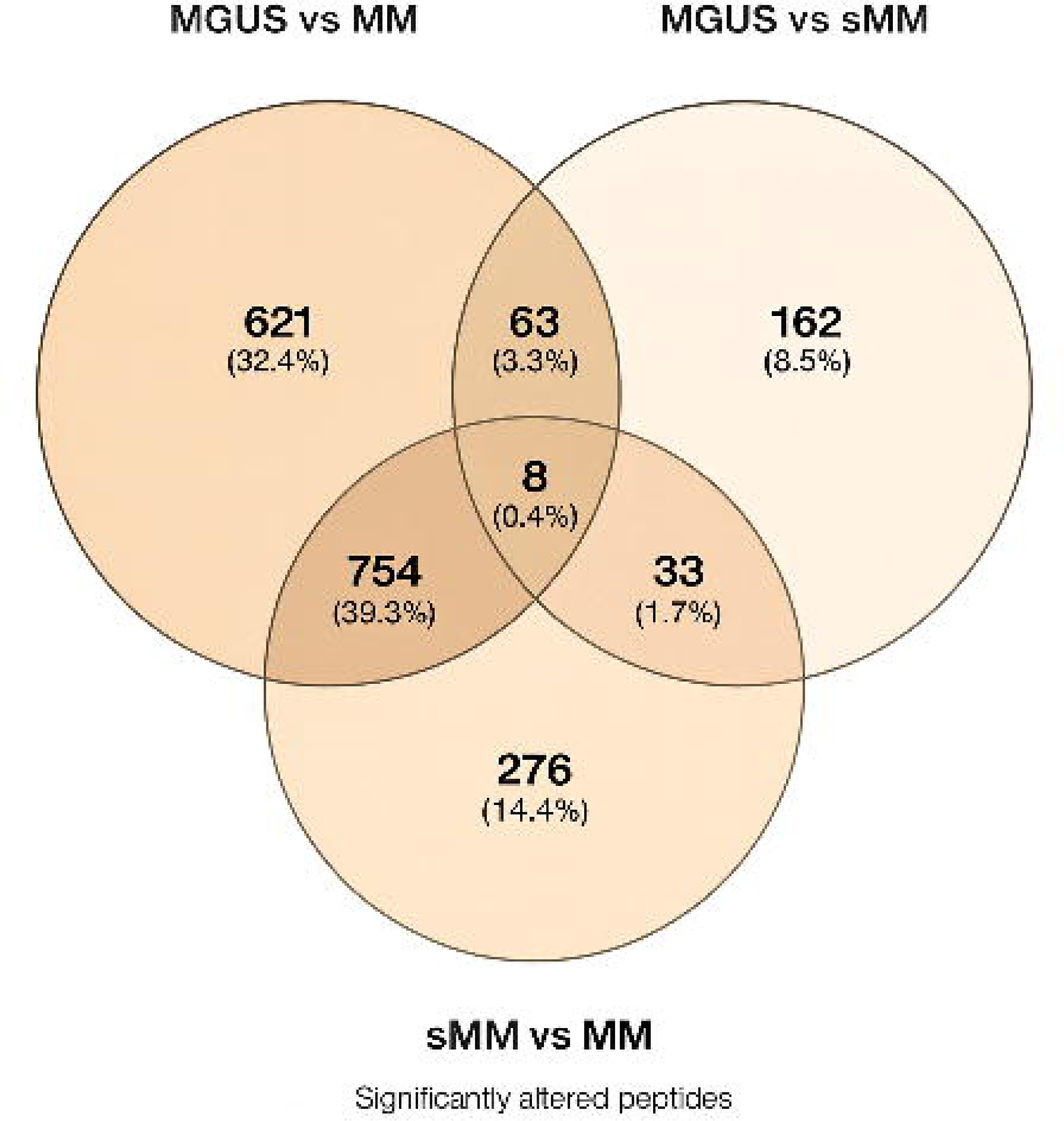
A) Commonly significant peptide biomarkers between three comparisons at 30% frequency threshold, and B) Heat map of the relative expression levels of the 121 significant peptides, showing in most cases monotonic abundance trends across MM development.

### A 121-peptide classifier (M121) can accurately detect MM

A support vector machine (SVM) model was developed using MosaCluster (v1.6.5) to assess the discriminatory capacity of the combined 121 MM-associated urinary peptides (named M121). Internal cross-validation of M121 within the discovery cohort achieved an overall classification accuracy of 96%, indicating robustness in-sample discrimination. Importantly, the mean M121 classifier ranks were 27.4 for MGUS, 46.8 for SMM, and 76.5 for MM, demonstrating a stage-dependent shift in classifier output consistent with monotonic proteomic remodeling along the MGUS to SMM to MM biological continuum **(Figure 3A).** Independent validation of M121 in 61 patients yielded an AUC of 0.94 (95% CI: 0.85–0.99, p < 0.0001),100% sensitivity, and 82% specificity at the optimal decision threshold (**Figure 3B**), confirming significant discrimination between active MM and precursor conditions without compromising false-positive control.

**Figure 3:**
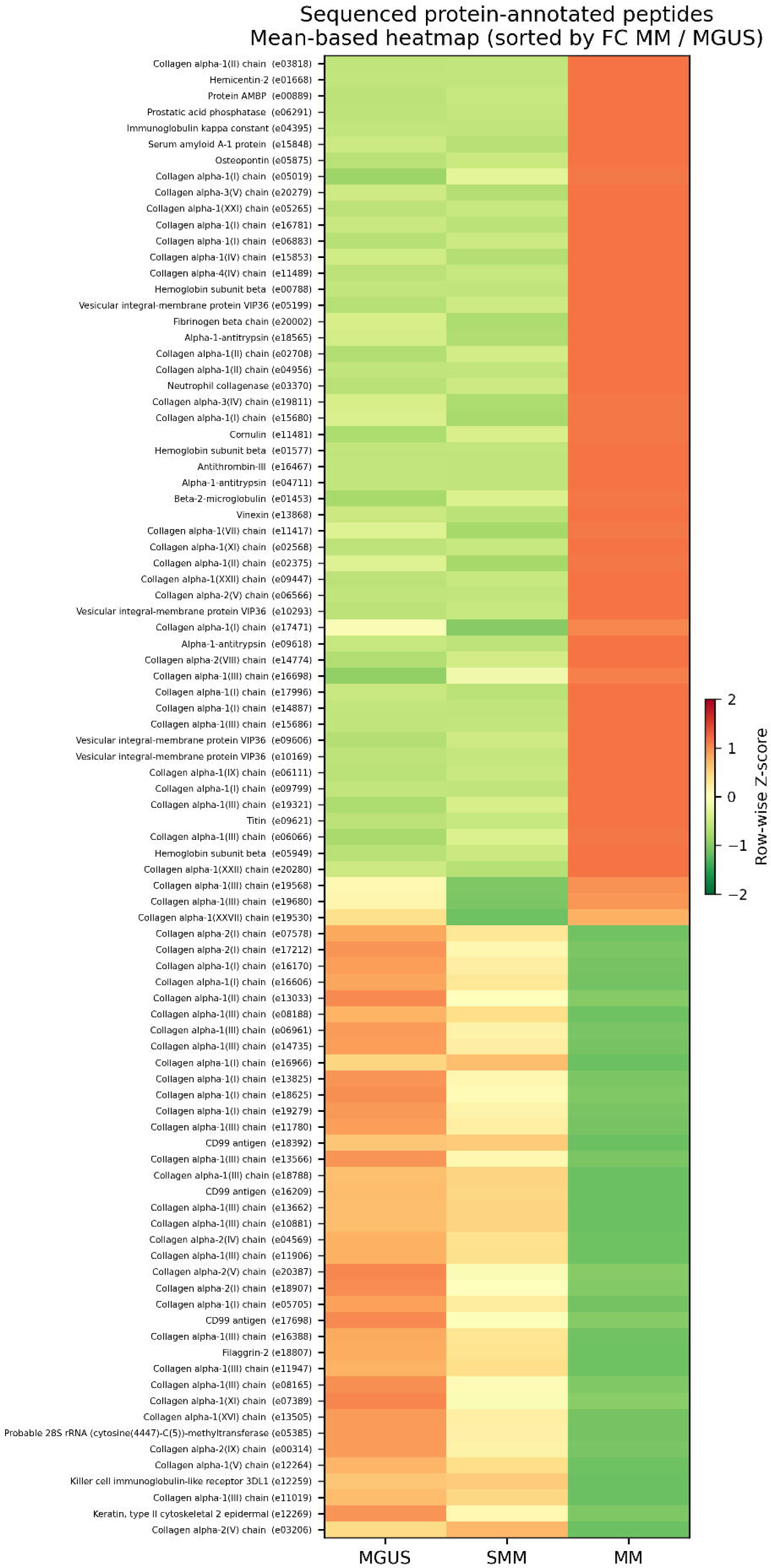
A) Classifier score distribution across MGUS, SMM, and MM showing stage-dependent monotonic increases consistent with progressive molecular transformation in the discovery cohort. B) Receiver operating characteristic (ROC) curve demonstrating performance of the M121 urinary peptide classifier in the validation cohort (AUC = 0.94).

### Assessment of the specificity and analytical robustness of the M121 classifier

To assess the M121 specificity, the classifier was applied to 86 urinary profiles from non-malignant reference individuals. All control samples yielded negative classification scores, confirming high specificity in healthy populations. The distribution of classifier scores across controls and MM cases is shown in the **Supplementary Figure S1.** The confounding effects of possible impaired kidney function were investigated by comparing the established classifier for MM (M121) with the well-established CE-MS-based classifier that detects CKD (namely CKD273), validated in multiple studies and clinical trials for early detection of CKD progression [24, 25]. A small overlap was observed at the single peptide level: eleven peptides were found common between M121 and the CKD273 classifier. To assess the specificity of the M121 at the SVM classification level, patients with known impaired kidney function (n=50) and controls from the same study [20] (patients at risk, but not developing AKI, n=25) were classified with M121. Ten of 75 patients (13.3%) scored positively using the M121 classifier, while the mean ranks based on M121 scoring were not significantly different between the patients at risk for AKI (35.5; n=25, 3 positive), those diagnosed with stage 3 AKI (38.7; n=25, 5 positive) and those with CKD (39.7; n=25; 2 positive p=0.7724). Analytical robustness was additionally evaluated across 196 repeated quality-control (QC) measurements obtained over multiple days. The coefficient of variation (CV) of the resulting classification score derived from CE–MS signal intensities was 5.6%, below the predefined 15% acceptance threshold **(Supplementary Figure S2).**

### Biological Characterization of MM-specific urinary peptides

Of the 121 MM-associated peptides, 92 were successfully sequenced and assigned to 41 parental proteins. The most frequently represented proteins included excretion of fibrillar and basement-membrane collagens (COL1A1, COL1A2, COL3A1, COL4A1), βL-microglobulin (B2M), α1-antitrypsin (SERPINA1), antithrombin-III (SERPINC1), CD99, serum amyloid A-1 (SAA1), and protein AMBP. Functional categorization indicated enrichment in ECM remodeling, immune regulation, inflammation, and plasma-cell adhesion, all central pathways in multiple myeloma pathogenesis. Several peptides demonstrated monotonic abundance changes consistent with disease evolution. In particular, urinary βL-microglobulin increased significantly from MGUS to MM, consistent with tumor burden–dependent proteolysis, whereas CD99 fragments showed progressive reduction along the same continuum. Protein-protein interaction (PPI) analysis using STRING (confidence ≥ 0.7) demonstrated significant network clustering (p < 1 × 10L¹L), connecting immune-regulatory proteins (e.g., B2M, SAA1) to fibrillar collagen hubs. Functional enrichment in Metascape revealed significant associations with the bone system (p = 2.1 × 10LL), connective tissue (p = 2.1 × 10LL), and plasma cells (although marginal; p = 0.038), supporting a biological origin in bone-marrow matrix degradation and plasma-cell dysregulation **(Figure 4).**

**Figure 4:**
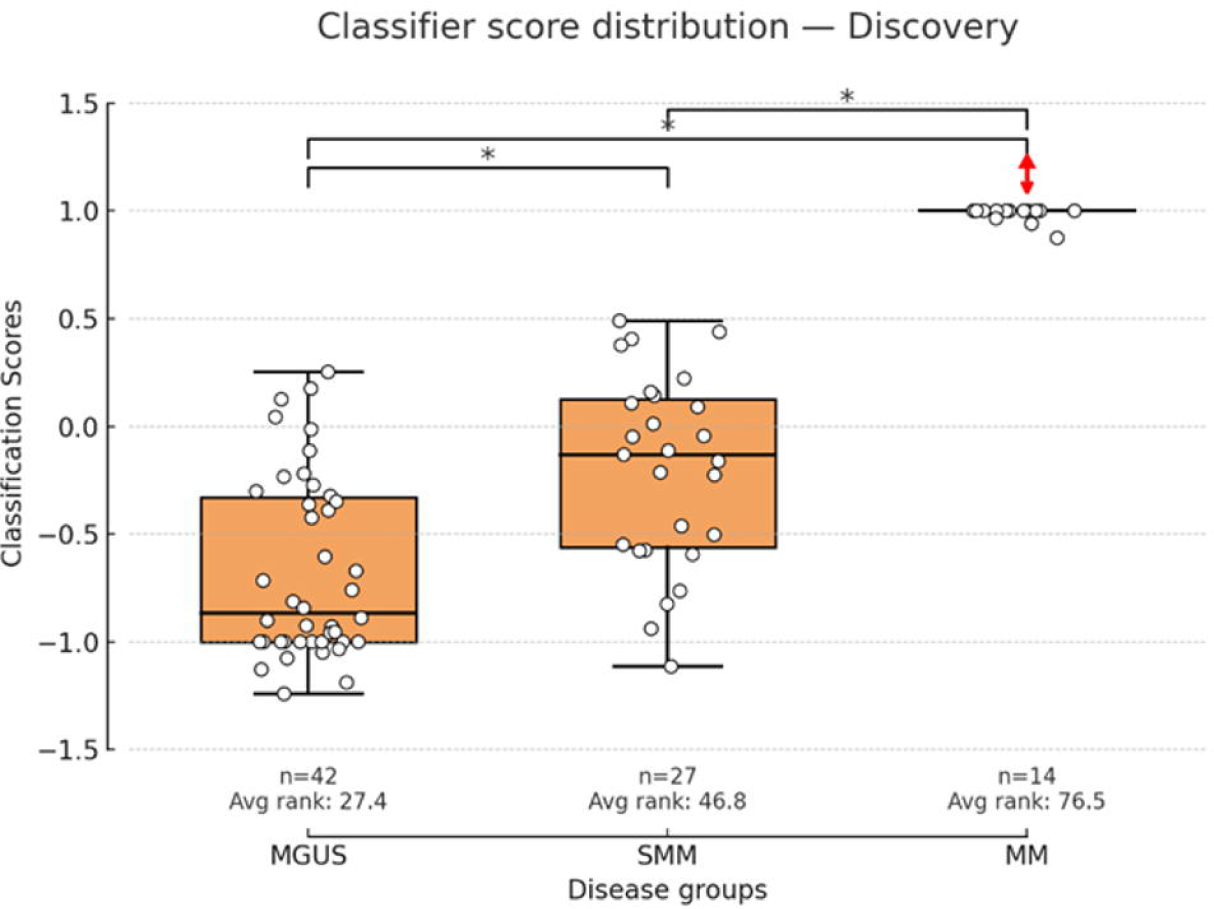
STRING protein–protein interaction (PPI) network of the 69 sequenced peptides (confidence ≥ 0.7). Immune mediators (orange nodes) cluster with collagen substrates (blue nodes), indicating coordinated remodeling of the extracellular matrix.

## Discussion

Following multiple studies on the application of CE-MS to profile ECM-related alterations reflected in the urinary peptidome in patients with solid tumors [26–28], in this cross-sectional feasibility study, we explored changes in urine peptides across the MGUS–SMM–MM disease spectrum. In plasma cell dyscrasias, where deterioration of renal function may contribute to detectable changes in the urinary peptidome, urinary peptidomics are especially relevant to investigate which signatures are specifically due to pathological proteolytic activity within the bone marrow microenvironment [19]. Using CE–MS–based urinary profiling, we developed and independently validated a 121-peptide classifier (M121) that discriminates active MM from precursor conditions with high accuracy and analytical reproducibility. The observed monotonic increase in classifier scores across MGUS, SMM, and MM suggests that the urinary peptide signature reflects, in parallel to tumor burden, dynamic stage-associated changes, likely in specific proteolytic activity. These findings extend current biomarker concepts in plasma-cell disorders, which remain primarily based on serological assays such as serum M-protein and free light-chain ratios, and bone-marrow plasma-cell infiltration [29, 30]. Conventional markers such as serum M-protein, involved/ uninvolved free light-chain ratio, and βL-microglobulin primarily quantify tumor burden, but do not directly capture microenvironmental remodeling processes [29, 31, 32]. Reported AUC values for βL-microglobulin and free light-chain ratio typically range between 0.72 and 0.78 in comparable MGUS–SMM–MM cohorts [33].

The progressive modulation of collagen-derived peptides, particularly those originating from collagen types I, III, and IV, highlights the contribution of ECM remodeling to myeloma pathophysiology. The stage-dependent increase in urinary collagen fragments reflects the osteolytic activity and proteolytic degradation occurring within the bone marrow microenvironment, driven by matrix metalloproteinases (MMPs), cathepsins, and inflammatory mediators [5, 34, 35]. Importantly, the systemic appearance of these peptides in urine indicates increased matrix turnover in the bone marrow but may also reflect broader connective-tissue remodeling [10, 36]. These observations are mechanistically aligned with recent proteomic and histopathologic studies showing that MMP- and cathepsin-mediated ECM degradation destabilizes the bone marrow niche and facilitates plasma-cell expansion and immune suppression [10]. Importantly, to assess potential confounding effects from impaired renal function present in MM patients, we compared the peptide signature with the well-established CE–MS-based CKD273 classifier, a known marker of kidney function and CKD progression. Eleven peptides were shared between the M121 classifier and CKD273. Exclusion of these peptides from the M121 model did not result in significant changes in classification scores (data not shown), indicating a stronger contribution of bone microenvironment remodeling rather than kidney function. This interpretation is further supported by the M121 classifier performance in patients with acute or chronic kidney disease. For the number of patients with impaired kindly function (AKI, or CKD) that were classified as positive using the M121 classifier, presence of an undiagnosed MGUS cannot be formally excluded as this was not systematically assessed in the AKI/CKD cohorts ; therefore, the However, the absence of significant differences in M121 scores across renal disease groups, together with the additional comparison with CKD273, suggests that the classifier signal is not primarily driven by renal impairment.In addition to collagen degradation products, several individual peptide markers showed biologically meaningful and stage-dependent regulation. Urinary βL-microglobulin increased progressively from MGUS to MM, consistent with its established use in Multiple Myeloma International Staging System (ISS) staging and tumor-burden estimation [2, 32]. Parallel increases in αL-antitrypsin and antithrombin-III fragments indicate inflammatory and coagulation pathway activation, in line with proteomic reports demonstrating systemic inflammation in MM progression [37]. Conversely, CD99-derived peptides decreased across advancing stages, supporting prior evidence that loss of CD99 in plasma cells promotes immune evasion and aggressive plasma-cell phenotypes, particularly in extramedullary disease [38]. Collectively, these coordinated peptide trends, in parallel to reflecting tumor mass also highlight remodeling of the ECM–immune regulatory axis as a central event in malignant transformation.

Technically, urinary sampling is non-invasive, and CE-MS is highly reproducible [27, 36, 39] while avoiding the spatial sampling bias associated with bone marrow biopsies [40]. Thus urinary peptidomics are suitable for repeated assessment, which needs to be proven in future longitudinal studies. The integration of 121 biologically driven peptide markers into an SVM model allows multidimensional representation of disease biology, capturing proteolytic remodeling processes that are not reflected in conventional single-analyte biomarkers [23, 41, 42]. From a translational perspective, urinary peptidomics may complement existing serum-based biomarkers by providing additional molecular information related to microenvironmental remodeling. In the present cross-sectional study, the observed stage-associated peptide patterns primarily support biological insights into disease-associated proteolytic processes. These data also present a solid basis for a longitudinal and multi-center investigation required to determine whether such peptide signatures have clinical utility beyond descriptive disease stratification[43, 44].

Despite these strengths, several limitations require consideration. First, the number of patients with overt multiple myeloma was small (n = 14 in discovery; n = 7 in validation), which may restrict the statistical generalizability of the classifier. The relatively limited number of MM cases reflects the pilot and hypothesis-generating design of the study and the stringent inclusion criteria applied. To address this limitation, model performance was assessed using multiple complementary metrics, including receiver operating characteristic analysis with confidence intervals and evaluation of classifier behavior across disease stages rather than reliance on binary classification alone. These measures were intended to support biological plausibility and reduce the likelihood of spurious associations arising from model complexity. Given the exploratory nature of this cross-sectional biomarker discovery and validation study, particular attention was paid to minimizing overfitting and ensuring robustness of the derived classifier. Feature selection, which was solely based on statistics and adjustment for multiple testing, was performed exclusively within the discovery cohort, and the M121 classifier development was followed by evaluation in an independent validation cohort that was not used at any stage of model training or optimization. Additionally, the observed monotonic expression of the classifier, together with its high specificity when applied in the general population cohort, enhances its validity. Moreover, only 92 of the 121 discriminatory peptides were successfully sequenced, leaving the biological identity of the remaining 29 peptides unresolved. Finally, the cross-sectional study design precludes assessment of whether M121 can prospectively predict progression from MGUS or smoldering myeloma. Future longitudinal studies, ideally with serial sampling, external multi-center validation, and integration with established risk models, will be essential and in fact, are being designed based on the data presented here to determine the prognostic utility and clinical adoption potential of this classifier.

## Conclusion

This study demonstrates that systemic ECM remodeling profiled by CE-MS can yield measurable features associated with advancing disease stages in the MGUS–SMM–MM spectrum. By integrating CE–MS urinary peptidomics with machine-learning classification, we developed and independently validated a 121-peptide urinary signature (M121) that distinguishes active multiple myeloma from precursor states with high analytical reproducibility in a fully non-invasive setting. Collectively, these findings support the feasibility of urinary peptidomics as a reproducible approach to study disease-associated microenvironmental remodeling in plasma-cell disorders.

## Associated Data

The data that support the findings of this study are openly available in Zenodo at https://doi.org/10.5281/zenodo.18094607

## Data Availability Statement

The proteomics data supporting the findings of this study are publicly available in Zenodo at https://doi.org/10.5281/zenodo.18094607

## Supporting information

Supplementary Table S1. CE-MS peptides to classify

## Acknowledgments

All authors are grateful to all patients who donated urine samples. This work was supported by funding through the European Union’s Horizon Europe ELMUMY (Grant Agreement No. 101097094) to EK, MAD, JZ, CL, MF, HM, and AV, funded by the European Commission. Views and opinions expressed are, however, those of the author(s) only and do not necessarily reflect those of the European Union. Neither the European Union nor the granting authorities can be held responsible for them. We also gratefully acknowledge the support of the Iran National Science Foundation (INSF) and the Ministry of Science, Research and Technology of Iran (MSRT). The authors would like to thank Prof. George Spyrou and Dr. Julie Courraud for their valuable discussions that contributed to the development of this work.

---

**Supplementary Figure 1:**
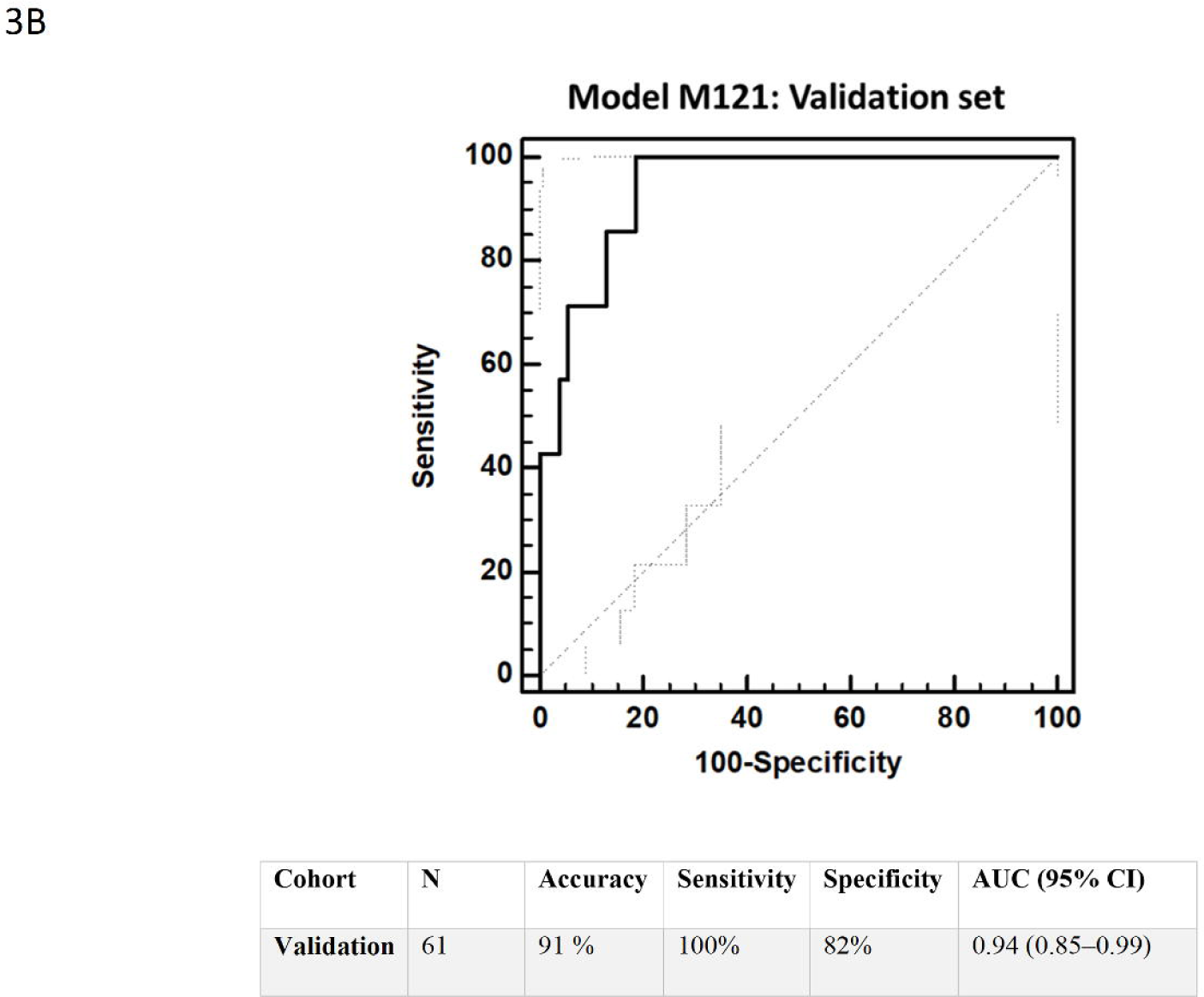
Classifier score distribution for M121 in non-malignant controls versus MM subjects, demonstrating complete separation and absence of false-positive classification.

**Supplementary Figure 2:**
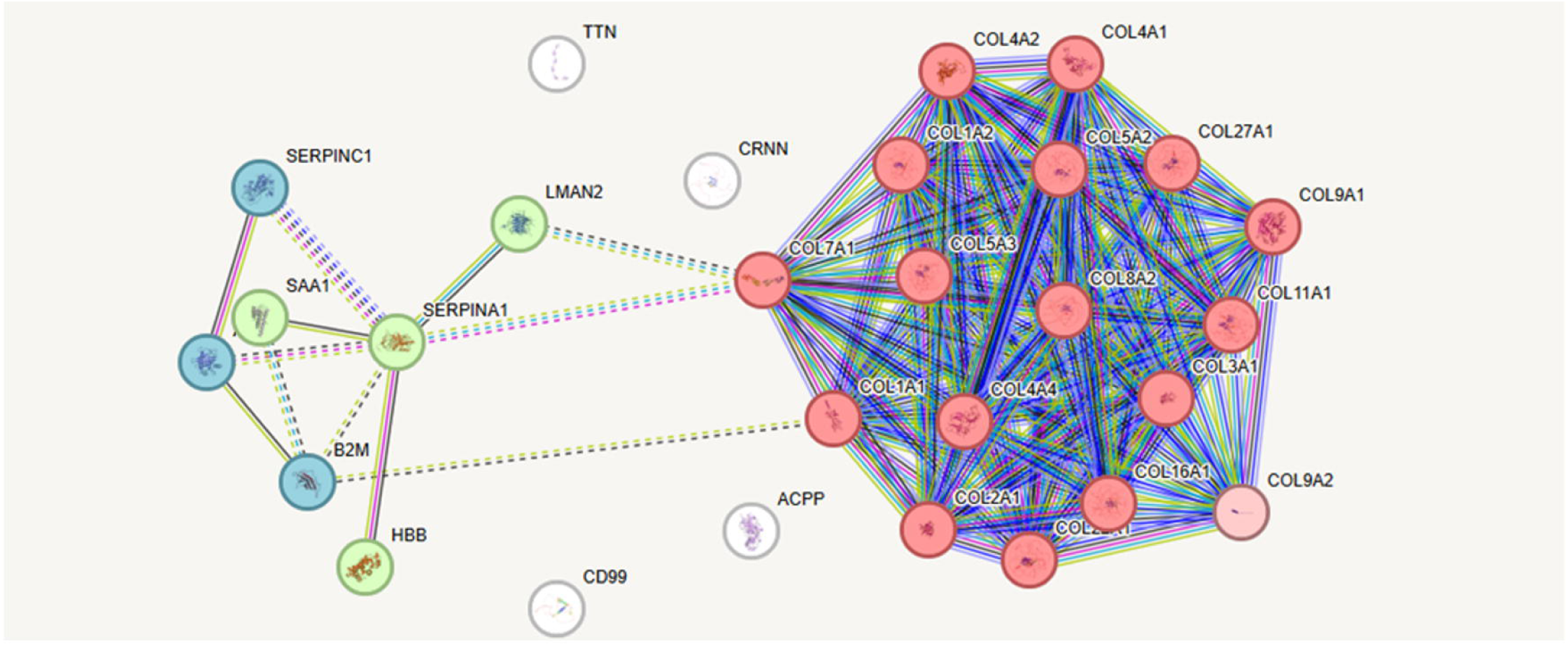
Analytical reproducibility of the classifier across 196 QC measurements, showing a coefficient of variation (CV) of 5.64 %.

## References

1. Abduh MS. (2024). An overview of multiple myeloma: A monoclonal plasma cell malignancy’s diagnosis, management, and treatment modalities. Saudi Journal of Biological Sciences 31(2), 103920.

2. Rajkumar SV. (2020). Multiple myeloma: 2020 update on diagnosis, risk-stratification and management. American journal of hematology 95(5), 548–67.

3. Testa U, Pelosi E, Castelli G, Leone G. (2024). Recent advances in the definition of the molecular alterations occurring in multiple myeloma. Mediterranean Journal of Hematology and Infectious Diseases 16(1), e2024062.

4. Kyle RA, Larson DR, Therneau TM, Dispenzieri A, Kumar S, Cerhan JR, et al. (2018). Long-term follow-up of monoclonal gammopathy of undetermined significance. New England journal of medicine 378(3), 241–9.

5. Naba A. (2024). Mechanisms of assembly and remodelling of the extracellular matrix. Nature Reviews Molecular Cell Biology 25(11), 865–85.

6. Neumeister P, Schulz E, Pansy K, Szmyra M, Deutsch AJ. (2022). Targeting the microenvironment for treating multiple myeloma. International Journal of Molecular Sciences 23(14), 7627.

7. Solimando AG, Da Vià MC, Bolli N, Steinbrunn T. (2022). The route of the malignant plasma cell in its survival niche: Exploring “Multiple myelomas”. Cancers 14(13), 3271.

8. Agafonova A, Prinzi C, Trovato Salinaro A, Ledda C, Cosentino A, Cambria MT, et al. (2025). Breaking barriers: the role of the bone marrow microenvironment in multiple myeloma progression. International Journal of Molecular Sciences 26(15), 7301.

9. Brassart-Pasco S, Brézillon S, Brassart B, Ramont L, Oudart J-B, Monboisse JC. (2020). Tumor microenvironment: extracellular matrix alterations influence tumor progression. Frontiers in oncology 10(397.

10. Socovich AM, Naba A, editors. The cancer matrisome: From comprehensive characterization to biomarker discovery. Seminars in cell & developmental biology; 2019: Elsevier.

11. Merz M, Hu Q, Merz AMA, Wang J, Hutson N, Rondeau C, et al. (2023). Spatiotemporal assessment of immunogenomic heterogeneity in multiple myeloma. Blood Advances 7(5), 718–33.

12. Mourad C, Cosentino A, Lalonde MN, Omoumi P, editors. Advances in bone marrow imaging: strengths and limitations from a clinical perspective. Seminars in musculoskeletal radiology; 2023: Thieme Medical Publishers, Inc.

13. Mavrogeorgis E, Mischak H, Latosinska A, Siwy J, Jankowski V, Jankowski J. (2021). Reproducibility evaluation of urinary peptide detection using CE-MS. Molecules 26(23), 7260.

14. Mischak H, Vlahou A, Ioannidis JP. (2013). Technical aspects and inter-laboratory variability in native peptide profiling: The CE–MS experience. Clinical biochemistry 46(6), 432–43.

15. Krochmal M, Schanstra JP, Mischak H. (2018). Urinary peptidomics in kidney disease and drug research. Expert opinion on drug discovery 13(3), 259–68.

16. Pontillo C, Mischak H. (2017). Urinary peptide-based classifier CKD273: towards clinical application in chronic kidney disease. Clinical kidney journal 10(2), 192–201.

17. Rajkumar SV, Dimopoulos MA, Palumbo A, Blade J, Merlini G, Mateos M-V, et al. (2014). International Myeloma Working Group updated criteria for the diagnosis of multiple myeloma. The lancet oncology 15(12), e538–e48.

18. Inker LA, Eneanya ND, Coresh J, Tighiouart H, Wang D, Sang Y, et al. (2021). New creatinine-and cystatin C–based equations to estimate GFR without race. New England Journal of Medicine 385(19), 1737–49.

19. Latosinska A, Siwy J, Mischak H, Frantzi M. (2019). Peptidomics and proteomics based on CE-MS as a robust tool in clinical application: The past, the present, and the future. Electrophoresis 40(18-19), 2294–308.

20. Piedrafita A, Siwy J, Klein J, Akkari A, Amaya-Garrido A, Mebazaa A, et al. (2022). A universal predictive and mechanistic urinary peptide signature in acute kidney injury. Critical Care 26(1), 344.

21. Crutchfield CA, Thomas SN, Sokoll LJ, Chan DW. (2016). Advances in mass spectrometry-based clinical biomarker discovery. Clinical proteomics 13(1), 1.

22. Jantos-Siwy J, Schiffer E, Brand K, Schumann G, Rossing K, Delles C, et al. (2009). Quantitative urinary proteome analysis for biomarker evaluation in chronic kidney disease. Journal of proteome research 8(1), 268–81.

23. Dimopoulos MA, Terpos E, Boccadoro M, Moreau P, Mateos M-V, Zweegman S, et al. (2025). EHA–EMN Evidence-Based Guidelines for diagnosis, treatment and follow-up of patients with multiple myeloma. Nature Reviews Clinical Oncology 22(9), 680–700.

24. Tofte N, Lindhardt M, Adamova K, Bakker SJ, Beige J, Beulens JW, et al. (2020). Early detection of diabetic kidney disease by urinary proteomics and subsequent intervention with spironolactone to delay progression (PRIORITY): a prospective observational study and embedded randomised placebo-controlled trial. The lancet Diabetes & endocrinology 8(4), 301–12.

25. Curovic VR, Eickhoff MK, Rönkkö T, Frimodt-Møller M, Hansen TW, Mischak H, et al. (2022). Dapagliflozin improves the urinary proteomic kidney-risk classifier CKD273 in type 2 diabetes with albuminuria: a randomized clinical trial. Diabetes Care 45(11), 2662–8.

26. Heidegger I, Frantzi M, Salcher S, Tymoszuk P, Martowicz A, Gomez-Gomez E, et al. (2024). Prediction of clinically significant prostate Cancer by a specific collagen-related Transcriptome, Proteome, and Urinome signature. European Urology Oncology

27. Belczacka I, Latosinska A, Siwy J, Metzger J, Merseburger AS, Mischak H, et al. (2018). Urinary CE-MS peptide marker pattern for detection of solid tumors. Scientific reports 8(1), 5227.

28. Mengual L, Frantzi M, Mokou M, Ingelmo-Torres M, Vlaming M, Merseburger AS, et al. (2022). Multicentric validation of diagnostic tests based on BC-116 and BC-106 urine peptide biomarkers for bladder cancer in two prospective cohorts of patients. British Journal of Cancer 127(11), 2043–51.

29. Rajkumar SV, Kumar S, Lonial S, Mateos MV. (2022). Smoldering multiple myeloma current treatment algorithms. Blood cancer journal 12(9), 129.

30. Went M, Duran-Lozano L, Halldorsson GH, Gunnell A, Ugidos-Damboriena N, Law P, et al. (2024). Deciphering the genetics and mechanisms of predisposition to multiple myeloma. Nature Communications 15(1), 6644.

31. Zanwar S, Rajkumar SV. (2025). Current risk stratification and staging of multiple myeloma and related clonal plasma cell disorders: MULTIPLE MYELOMA, GAMMOPATHIES. Leukemia 1–8.

32. Murray D, Dispenzieri A, Kumar S, Gill H, Vachon C, Snyder M, et al. (2020). Free light chain assay drift: potential for misdiagnosis? The journal of applied laboratory medicine 5(6), 1411–3.

33. Dispenzieri A, Kyle RA, Katzmann JA, Therneau TM, Larson D, Benson J, et al. (2008). Immunoglobulin free light chain ratio is an independent risk factor for progression of smoldering (asymptomatic) multiple myeloma. Blood, The Journal of the American Society of Hematology 111(2), 785–9.

34. Prakash J, Shaked Y. (2024). The interplay between extracellular matrix remodeling and cancer therapeutics. Cancer discovery 14(8), 1375–88.

35. Frantzi M, Latosinska A, Belczacka I, Mischak H. (2019). Urinary proteomic biomarkers in oncology: ready for implementation? Expert review of proteomics 16(1), 49–63.

36. Klein J, Bascands J-L, Mischak H, Schanstra JP. (2016). The role of urinary peptidomics in kidney disease research. Kidney international 89(3), 539–45.

37. Morè S, Corvatta L, Manieri VM, Morsia E, Offidani M. (2024). The challenging approach to multiple myeloma: From disease diagnosis and monitoring to complications management. Cancers 16(12), 2263.

38. Roshal M, Gao Q, Hutcherson S, Thoren K, Zhu M, Murata K. (2024). Evaluation of Plasma Cell Neoplasms. Manual of Molecular and Clinical Laboratory Immunology 2(1206-23.

39. Mokou M, Lygirou V, Vlahou A, Mischak H. (2017). Proteomics in cardiovascular disease: recent progress and clinical implication and implementation. Expert review of proteomics 14(2), 117–36.

40. Shama A, Soni T, Jawanda IK, Upadhyay G, Sharma A, Prabha V. (2023). The latest developments in using proteomic biomarkers from urine and serum for non-invasive disease diagnosis and prognosis. Biomarker Insights 18(11772719231190218.

41. Vapnik V. The nature of statistical learning theory: Springer science & business media; 2013.

42. Gadalla AA, Friberg IM, Kift-Morgan A, Zhang J, Eberl M, Topley N, et al. (2019). Identification of clinical and urine biomarkers for uncomplicated urinary tract infection using machine learning algorithms. Scientific reports 9(1), 19694.

43. Dunphy K, O’Mahoney K, Dowling P, O’Gorman P, Bazou D. (2021). Clinical proteomics of biofluids in haematological malignancies. International Journal of Molecular Sciences 22(15), 8021.

44. Ahangar M, Mischak H, Moulavasilis N, Stravodimos K, Mahjoubi F, Jankowski J, et al. (2025). Urinary peptidomic signatures predict overall and progression-free survival in patients with bladder cancer. medRxiv 2025.10. 28.25338944.

