## Supplementary Figures for "Peptidomic profiling reveals extracellular matrix remodeling signatures discriminative of multiple myeloma"

***Supplementary Figure 1***

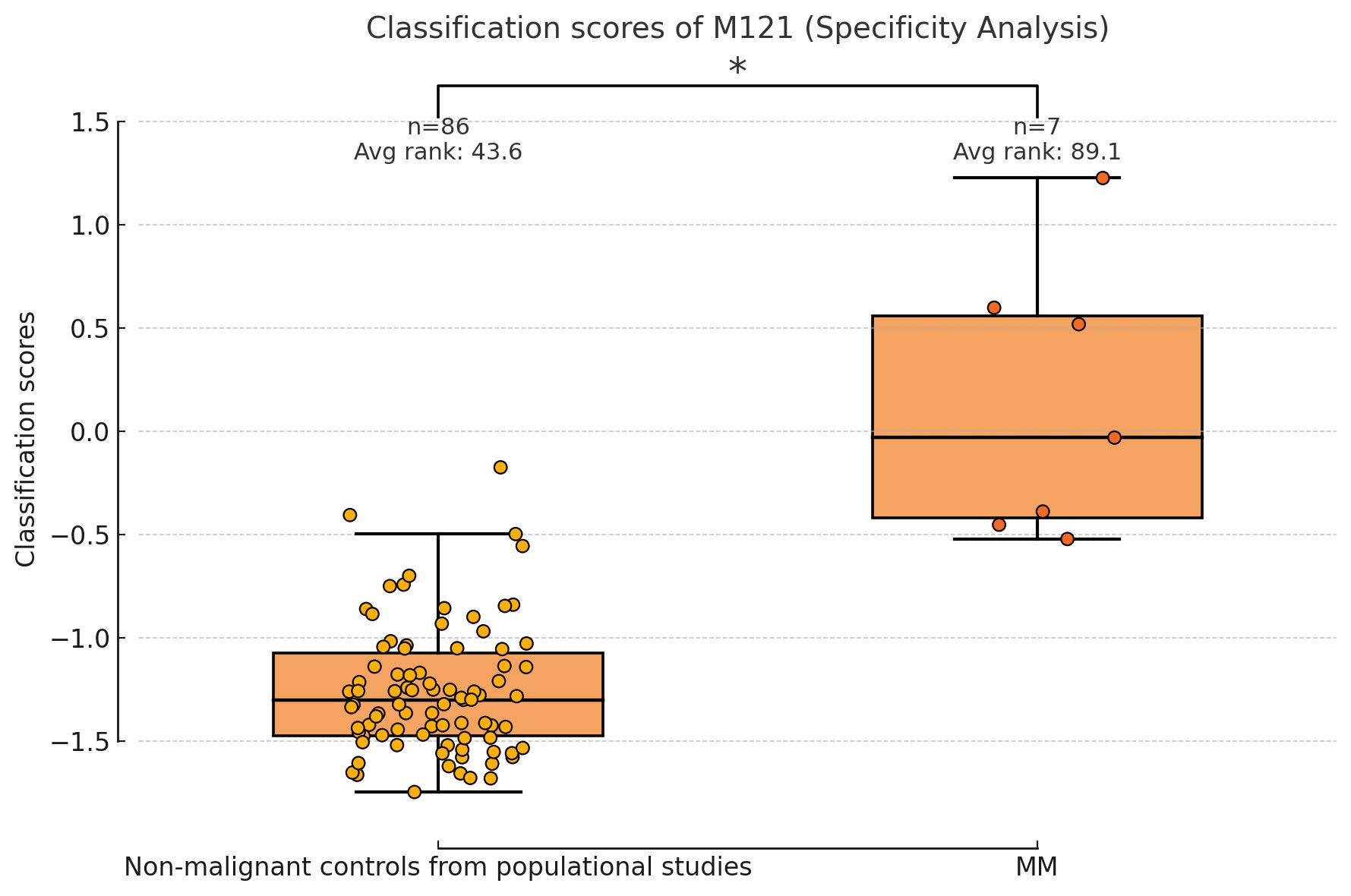

***Supplementary Figure 1:*** *Classifier score distribution for M121 in non-malignant controls versus MM subjects, demonstrating complete separation and absence of false-positive classification.*

***Supplementary Figure 2***

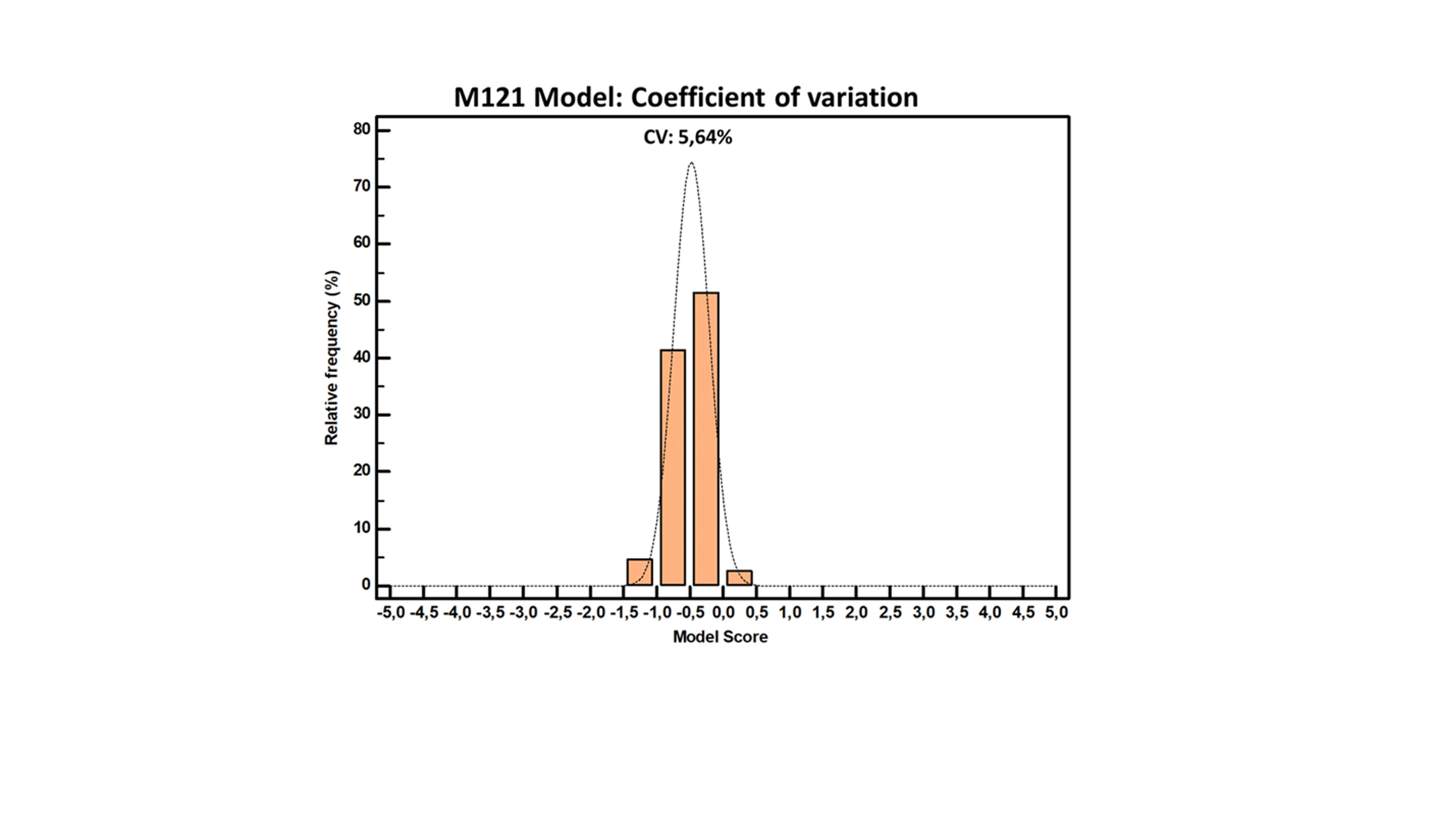

***Supplementary Figure 2:*** *Analytical reproducibility of the classifier across 196 QC measurements, showing a coefficient of variation (CV) of 5.64 %.*
